# Identification of clinical features associated with mortality in COVID-19 patients

**DOI:** 10.1101/2021.04.19.21255715

**Authors:** Rahimeh Eskandarian, Zahra Alizadeh Sani, Mohaddeseh Behjati, Mehrdad Zahmatkesh, Azadeh Haddadi, Kourosh Kakhi, Mohamad Roshanzamir, Afshin Shoeibi, Roohallah Alizadehsani, Sadiq Hussain, Fahime Khozeimeh, Vahideh Keyvani, Abbas Khosravi, Saeid Nahavandi, Sheikh Mohammed Shariful Islam

## Abstract

**Background:** To prevent infectious diseases, it is necessary to understand how they are spread and their clinical features. Early identification of risk factors and clinical features is needed to identify critically ill patients, provide suitable treatments, and prevent mortality.

**Methods:** We conducted a prospective study on COVID-19 patients referred to a tertiary hospital in Iran between March and November 2020. Of the 3008 patients (mean age 59.3±18.7 years, range 1 to 100 years), 1324 were women. We investigated COVID-19 related mortality and its association with clinical features including headache, chest pain, symptoms on CT, hospitalization, time to infection, history of neurological disorders, having a single or multiple risk factors, fever, myalgia, dizziness, seizure, abdominal pain, nausea, vomiting, diarrhoea and anorexia.

**Findings:** There was a significant association between COVID-19 mortality and old age, headache, chest pain, respiratory distress, low respiratory rate, oxygen saturation less than 93%, need for a mechanical ventilator, having symptoms on CT, hospitalization, time to infection, history of hypertension, neurological disorders, cardiovascular diseases and having a risk factor or multiple risk factors. In contrast, there was no significant association between mortality and gender, fever, myalgia, dizziness, seizure, abdominal pain, nausea, vomiting, diarrhoea and anorexia.

**Interpretation:** Our results might help identify early symptoms related to COVID-19 and better manage patients clinically.

## 1. Introduction

In January 2020, severe acute respiratory syndrome coronavirus 2 (SARS-CoV-2), was discovered [1]. Since then, the virus has spread exponentially and caused immense human sufferings worldwide [2-4]. The high number of deaths and the global spread of coronavirus disease (COVID-19) led the World Health Organization to announce it as a pandemic on 12 March 2020. The world has suffered a high toll from this pandemic regarding increased poverty, economic repercussions and human lives lost to date. A considerable portion of the population being asymptomatic carriers for COVID-19. Fever (83%), cough (82%) and shortness of breath (31%) are reported as the most common symptoms [5]. Chest X-ray generally demonstrates ground-glass opacity and multiple mottling in patients with pneumonia. COVID-19 patients typically yield decreased eosinophils and lymphocyte counts, lower median haemoglobin values, and enhance neutrophil counts, WBC and serum levels of ALT, AST, LDH and CRP [6]. For severe COVID-19 development, initial CRP serum levels have been considered an independent predictor [7]. Although the lung is the main target of COVID-19 infection, the widespread distribution of ACE2 receptors in organs [8] may lead to gastrointestinal, liver, kidney, central nervous system, cardiovascular and ocular damage needs to be closely observed [9]. Patients with acute respiratory distress syndrome may deteriorate speedily and die of multiple organ failure [5] induced by the so-called “cytokine storm”. The severity of COVID-19 is also associated with elevation of D-dimer levels. The elevated D-dimer levels may reflect the risk of disseminated coagulopathy in patients with severe COVID-19, which may require anticoagulant therapy [10].

#### Research in context

##### Evidence before this study

To prevent infectious diseases, it is necessary to understand how they are spread and their clinical features. Early identification of risk factors and clinical features is needed to identify critically ill patients, provide suitable treatments, and prevent mortality. As the best of our knowledge, a few researchers focused in investigation of important factors of mortality rate in COVID-19. Consequently, a few features have been investigated precisely.

##### Added value of this study

As far as we know, there is a few papers published in this field. In this research, the influence of different factors in COVID-19 patient’s death were investigated. In addition, there are some features which we investigated for the first time.

##### Implications of all the available evidence

Findings these factors could help physicians to better setting the priority of treatments for patients. It results to death rate reduction. By identifying patients at risk, physicians can ascertain those who might benefit from early monitoring.

Early surveillance, contact tracing, testing, and strict quarantine are the strategies of the countries that had maintained a low COVID-19 mortality rate [11, 12]. Such countries had adopted digital technology to implement effective strategies and integrating them with healthcare. Pandemic plans are thorny to achieve manually but can be facilitated using digital health technology. Early flattening of the incidence curve was possible for some countries like South Korea, which had integrated government-coordinated mitigation and containment processes into digital technology [13]. The tracking of COVID-19 had initiated AI dashboards that showcased the disease burden. UpCodeto utilizes the data generated by the Singapore Ministry of Health to portray infection trends and recovery time [14]. The web-based platform HealthMap and COVID-19 dashboard of the Johns Hopkins University provide an up-to-date scenario of COVID-19 deaths and cases across the world [15]. AI algorithms play a vital role in the integration of digital technology with healthcare. But AI algorithms have constraints as they require a large amount of data to provide accurate results [16]. Most of the predictive COVID-19 models were based on Chinese data; hence it might not help other parts of the world. Social media and other online traffic and the absence of historical training data created noise in big datasets and produced overfitted models. These noises must be filtered to discern accurate predictions and trends. To study the risk factors, characteristics, and clinical outcome associated with COVID-19 mortality and other important factors.

Shi et al. [17] analysed the characteristics, risk factors and outcomes for in-hospital mortality of COVID-19 patients with diabetes. They abstracted laboratory, clinical, and demographic data of the patients and the risk factors associated with mortality were identified by performing Multivariable Cox regression analyses. The outcomes of COVID-19 patients with diabetes were lower than age- and sex-matched patients without diabetes.

Ji, et al. [18] exploited the association between healthcare resource availability and COVID-19 mortality. The comprehensive analysis of data from China showcased substantial disparities in mortality rates among various regions of Hubei (about 2·9% on average), Wuhan (>3%), and across the other provinces of China (on average about 0·7%). The empirical results demonstrated a striking positive correlation yielding healthcare burden was correlated with mortality. The coastal provinces of China, like Guangdong and Zhejiane, were low mortality rates despite having a high number of patients proved the point. Li et al. [19] evaluated the severity of the admission of treatment, complications and outcomes of patients with COVID-19. A multivariable binary logistic model was devised to analyse the vital risk factors for severe COVID-19 patients. They observed that severe male patients with hyperglycemia, heart injury, and high-dose corticosteroid use might risk mortality higher.

Selenium (Se) is a crucial trace element for human health and needed for a balanced immune response. The authors [20] hypothesized that Se status was correlated with the death of COVID-19 patients. They showed that Se status was considerably lower in samples from non-survivors as compared to surviving COVID patients. Hence, the role of Se for COVID-19 patients was strengthened by their findings. Qin et al. [21] carried out a retrospective study to assess the prognostic power and associations of circulating cardiac injury markers with the severe outcome of COVID-19 patients. They concluded that COVID-19 patients with elevated cardiac injury markers above the novel recognized cutoffs were associated with an elevated risk of mortality. The current reference standards showed higher prognostic cutoff values than these biomarkers.

Statins are recommended as an adjunct therapy for COVID-19 and are lipid-lowering therapeutics with favourable anti-inflammatory profiles. In [22], the dataset of 13,981 patients with COVID-19 from China was explored, out of which 1,219 received statins. Marginal structural and Cox time-varying model analyses were utilized to observe the statin use-associated lower risk of mortality. They asserted that to combat the mortality of COVID-19 patients applying statin treatment required further study and validation. The nutrition status of COVID-19 patients is not known. Zhao et al. [23] examined critically and severely ill COVID-19 patients’ nutrition and clinical characteristics and evaluated the relationship between clinical outcomes and nutrition risk. They demonstrated that 1-unit elevation in NRS scores was related to a more extended hospital stay and a higher risk of mortality in their logistic regression model.

Yadaw et al. [24] devised a useful prediction model of COVID-19 mortality utilizing unbiased computational techniques and detected the most predictive clinical features. Their machine learning framework was mainly based on three clinical features: minimum oxygen saturation throughout patients’ medical encounters, age, and type of patient encounter. Their COVID-19 mortality prediction model exhibited a competitive accuracy.

It is a hot research area whether the severity of COVID-19 health outcomes, including death, is associated with long-term exposure to air pollution. But the limitation of COVID-19 data availability and quality hinders in conducting decisive studies in this regard. COVID-19 outcome publicly available data for representative populations are available as area-level counts. Hence, In [25], an ecological regression analysis was studied that precluded controlling for individual-level COVID-19 risk factors.

Zhou et al. [26] exploited the clinical, demographic, and laboratory data, including serial samples for viral RNA detection from the electronic health records from hospitals in China and compared between non-survivors and survivors. They utilized univariable and multivariable logistic regression methods to study the risk factors associated with in-hospital mortality. The potential risk factors of the high SOFA score, older age, and d dimer greater than 1 μg/mL could help clinicians diagnose patients with poor prognosis at an early stage. In Supplementary Table S2, the relationship between some of the common clinical features of COVID-19 patients with mortality rate is listed. Although a number of studies have explored the association of mortality with clinical features of COVID-19, those studies did not provide a comprehensive list of list clinical features associated with COVID-19 mortality. In this study, we aimed to determine the association of a wide range of clinical features associated with COVID-19 mortality for the first time.

## 2. Method

From March 2020 to November 2020, 3008 patients with COVID-19, of which 94.5% (2844 cases) were of Iranian nationality and 5.5% (164 cases) were Afghan nationals, were examined in this experiment. Of these 3008 cases infected with COVID-19, 56% were men, and 44% were women with an age average of 59.3±18.7 years (1-100 years). In Figure 1, the number of patients in a different range of ages and their mortality numb is illustrated. Of the patients admitted to the hospital during this period, 18.5% required to be admitted to the intensive care unit and the rest to the isolated and normal wards. 387 patients (12.9%) with Covid-19 were in contact with the infected person, and 2621 patients (87.1%) declared any contact with the infected person. About 70.4% of patients referred us personally, and 653 (21.7%) of them conveyed to the hospital by pre-hospital emergency, 199 (6.6%) by private ambulance and 38 (1.3%) by ambulances available in other centres.

**Figure 1.**
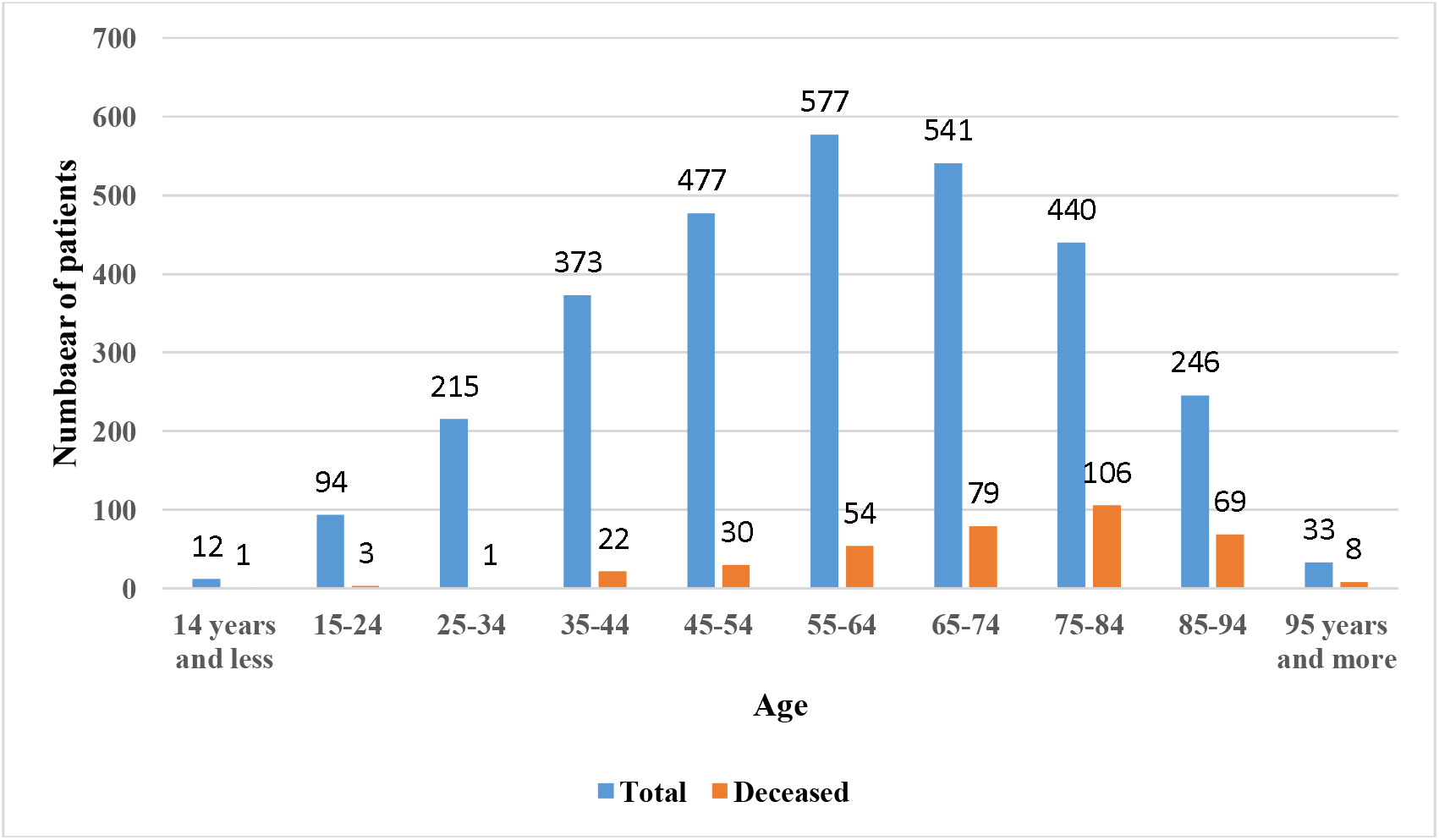
The relationship between mortality and age

PCR test was performed for 2822 cases (93.8%), of which 1431 cases (47.6%) were positive, and 1286 cases (42.8%) were negative. PCR test was not performed in 186 patients (6.2%) and was not reported for 104 patients (3.5%). Of the studied patients, 20 patients (0.7%) had a history of the previous infection. Patients admitted to the hospital were associated with symptoms including 32.2% fever, 28% cough, 14% myalgia, 43.3% loss of consciousness, 0.8% loss of sense of smell, 0.5% loss of taste, 0.4% seizures, 4.6% headache, 1.6% dizziness, 0.4% paresis, 0.1% plague, 3.8% chest pain, 3.8% chills, 0.5% sweating, 0.5% dry throat and sore throat, 7.8% weakness and lethargy, 0.2% sputum excretion, 0.2% gastrointestinal bleeding, 2.3% abdominal pain, 5.4% Nausea, 3.8% vomiting, 2.9% diarrhoea and 4.4% anorexia. Other initial symptoms included hemoptysis (in 2 patients), edema, restlessness, delirium, earache, constipation, palpitations, sudden loss of vision, and hematuria (each in one case). Fifty cases (1.7%) were a smoker, and 70 cases (2.3%) were addicted to drugs. 2764 patients underwent CT scan, of which 2277 had symptoms, and 244 did not undergo CT scan. 178 patients (5.9%) needed mechanical ventilation at the beginning of the study, and the others did not. The average (± standard deviation) level of oxygen saturation at referral was 89.3%±7.4% (39%-100%). 37.2% of patients had more than 93% oxygen saturation.

The number of patients’ respiration per minute were also measured in such a way that 0.3% (9 patients) did not breathe at all, 194 patients (6.4%) with 10-14 breaths, 1068 patients (35.5%) had 14-18 breaths and 1296 patients (43.1%) showed 18 122 breaths per minute. Indeed, 353 patients (11.8%) had 22-28 breaths, and 88 patients (2.9%) had more than 28 breaths per minute. The average (±SD) of patients’ body temperature at the time of referral was 37.1 ± 0 0.7 °C (35-40). 21.8% of patients had a fever at the time of referral.

The average (± SD) duration of symptoms until referral was 4.7± 13.9 days. In these patients, 1670 patients (55.5%) had risk factors or underlying diseases, so that 104 patients (3.5%) had cancer, 16 patients (0.5%) had liver disease, 588 patients (19.5%) with diabetes, 39 (1.3%) with chronic haematological diseases, 15 (0.5%) with immunodeficiency, 586 patients (19.5%) with cardiovascular diseases, 177 patients (5.9%) with kidney diseases, 108 patients (3.6%) with asthma, 99 patients (3.3%) with chronic lung diseases, 127 patients (4.2%) with neurological diseases, 695 patients (23.1%) with hypertension, 26 patients (0.8%) with CVA and stroke, 8 patients (0.2%) with neurosurgery related problems, 28 patients (0.9%) with hypothyroidism, 43 patients (1.4%) with other neurological diseases, 42 patients (1.3%) with hyperlipidemia, 15 patients (0.4%) with prostate, 43 patients (1.4%) with psychological diseases, 10 patients (0.3%) with history of veteran chemical warfare and 24 patients (0.7%) had anemia. Among them, 6 cases (0.5%) were pregnant. Out of 177 patients with kidney disease, 77 were on dialysis.

1338 patients (44.5%) had no risk factor and underlying disease. 823 (27.4%) and 567 (18.8%) patients had one and two risk factors, respectively. Three risk factors were observed in 218 (7.2%) and 52 cases (1.7%), respectively. Nine (0.3%) and one patient (0.05%) had five and six risk factors, respectively. Of these patients, 112 (3.7%) were hospitalized, 2523 (83.9%) were discharged, and also 373 (12.4%) died. The average (±SD) duration of hospitalization was 6.17 ± 6.3 days (1-87 days), of which 236 patients (7.8%) did not need hospitalization, and 2154 patients (71.6%) required 1-7 days of hospitalization. 376 cases (12.5%) 8-14 days, 137 cases (4.6%) 15-21 days, 59 cases (2%) 22-28 days and 16 cases (0.5%) more than 28 days were hospitalized.

According to these data, the prevalence of COVID-19 infection was high in March 2020 and then had the lowest incidence in May and June, and finally reached its peak in October and was associated with the lowest incidence in November (Figure 2).

**Figure 2.**
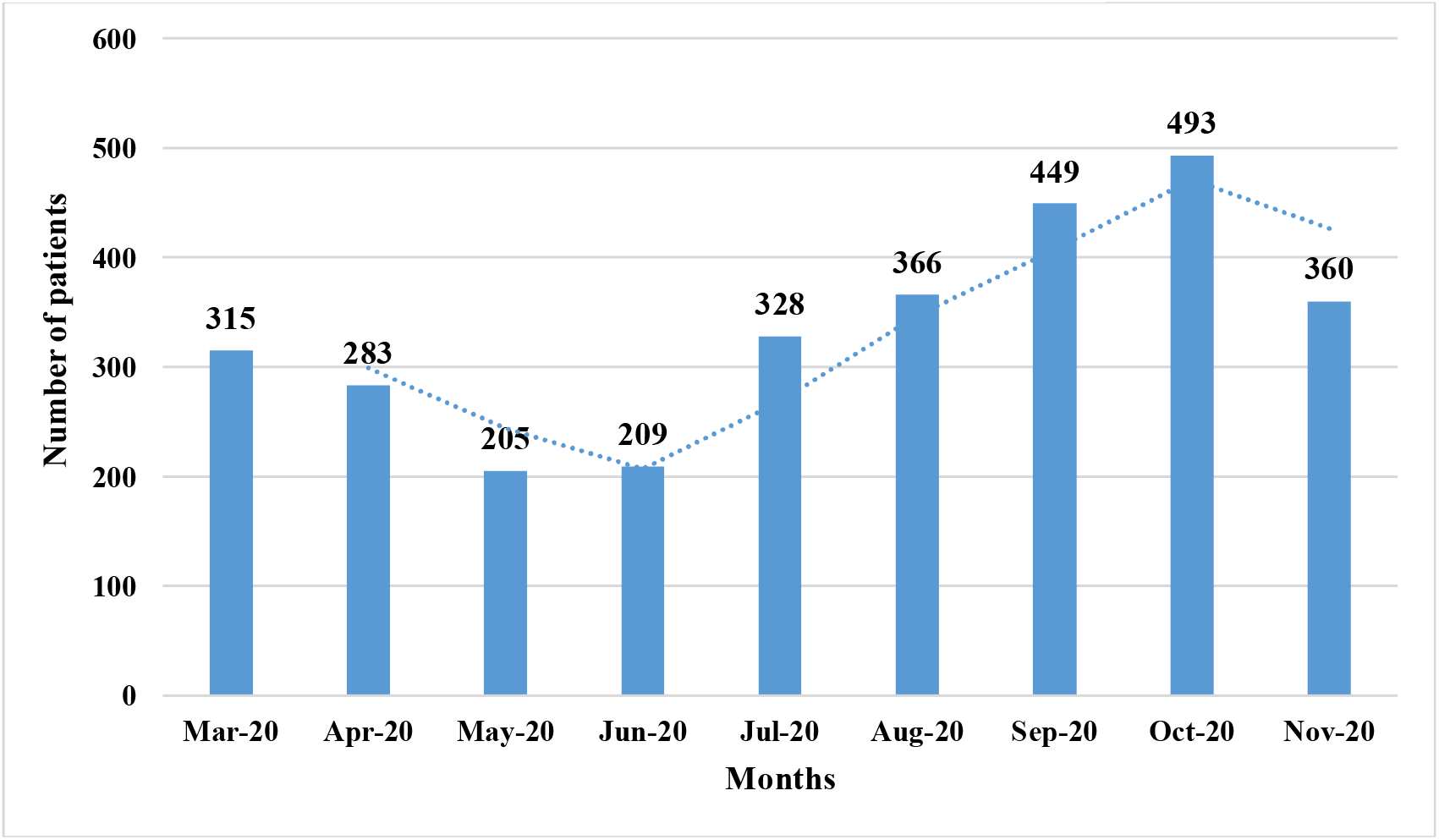
Number of patients between March 2020 and November 2020

Local ethical committee of the university approved this research. The patients were informed about this research aims, and written consent was obtained before data collection.

### Statistical analysis

Data are presented as the number of patients in a specific category and the mortality rate in each of them. We analysed the features using Matlab 2018b software. To specify differences between the two groups, Wilcoxon Rank□Sum Test 18 and Fisher’s exact test 17 were used for continuous and categorical data respectively. P ≤ .05 sets as the statistical significance.

## 3. RESULT

### The effect of early symptoms on the outcome of patients’ deaths

Table 1 shows the effect of different features on the mortality rate. Mortality was not significantly different between men (1684 cases) vs. women (1324 cases). There was a significant difference between mortality and age of patients (P <0.001), infection time (P <0.001), and the hospitalization ward (isolated ward, intensive care unit, normal ward) (P<0.001). Symptoms such as fever, myalgia, dizziness, seizure, abdominal pain, nausea, vomiting, diarrhoea and anorexia were occurred without having mortality related to COVID-19 (P>0.05). There was a significant difference between mortality and headache in patients infected with COVID-19 (P<0.011). Presentation of chest pain was also associated significantly with COVID-19 related mortality (P<0.045). Decreased level of consciousness was also significantly associated with COVID-19 related mortality (P<0.000). Respiratory distress, oxygen saturation less than 93%, lower respiratory rate and need for mechanical ventilation were associated with COVID-19 related mortality (P<0.004, P<0.000, P<0.000 and P<0.000, respectively).

**Table 1.**
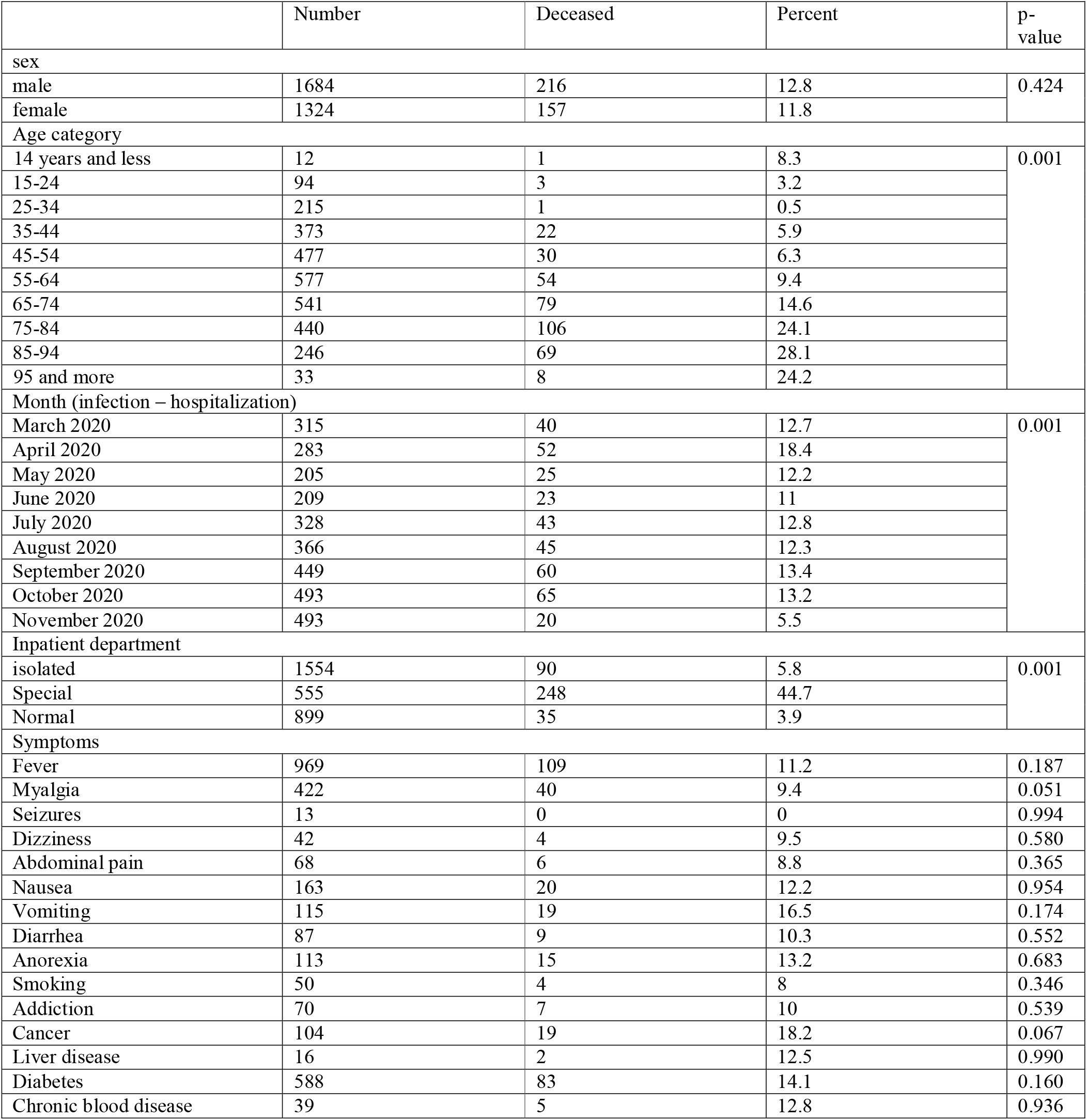
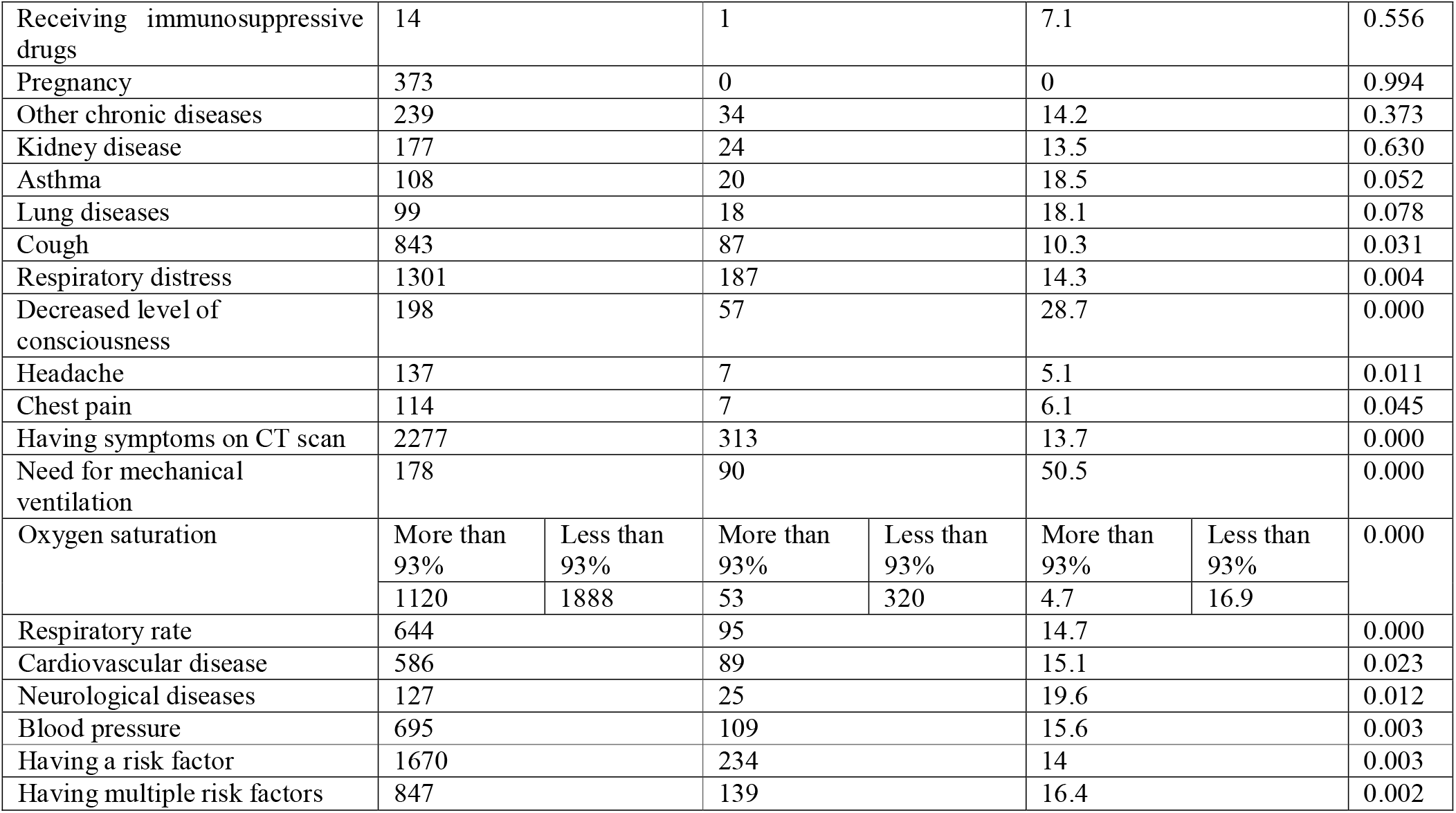
The effect of some features on the mortality rate

Opium addiction, smoking status, pregnancy, diabetes mellitus, underlying cancer, liver disease, lung disease, asthma, kidney disease, chronic haematological diseases, other chronic diseases, and receiving immunosuppressive medicines had no association COVID-19 related mortality. Underlying cardiovascular disease, hypertension and neurological diseases were associated with COVID-19 related mortality (P<0.023, P<0.003 and P<0.012, respectively). The presence of CT scan symptoms was significantly related to mortality in COVID-19 cases (P<0.000). There was a significant difference between mortality and having a risk factor for patients with COVID-19 (P < 0.003). There was a significant difference between mortality and the number of risk factors in patients with COVID-19 (P <0.002).

## 4. Discussion

The main findings of our study are the significant association between mortality of COVID-19 and old age, headache, chest pain, respiratory distress, low respiratory rate, oxygen saturation less than 93%, need to a mechanical ventilator, having symptoms on CT, hospitalization in wards, and time to infection. Besides, history of hypertension, neurological disorders, cardiovascular diseases, having a risk factor or multiple risk factors were associated with the COVID-19 mortality. Interestingly, there was no significant association between mortality and gender, fever, myalgia, dizziness, seizure, abdominal pain, nausea, vomiting, diarrhoea, and anorexia. We observed no association regarding COVID-19 related mortality and headache, chest pain, having symptoms on CT, hospitalization in wards and time to infection. Indeed, history of neurological disorders, having a risk factor or multiple risk factors, fever, myalgia, dizziness, seizure, abdominal pain, nausea, vomiting, diarrhoea and anorexia with COVID-19 related mortality were not found in other investigations.

The significant association between age and COVID-19 related mortality in our study is in line with previous studies conducted by Zhou et al. [26], Pettit et al. [27], Chen et al.[28], Iftime et al. [29], and in contrast to De Smet et al. [30], Sun et al. [31] and Li et al. [19]. Immune impairment and the enhanced possibility of developing cardiovascular and respiratory diseases would be the joint linkage between old age and COVID-19 related mortality [32]. The observed association between the underlying cardiovascular diseases and COVID-19 related mortality in our study was in line with Chen et al. [33], Soares et al.[34] and Ruan et al. [35], but was contrary to Iftimie et al. [36], Li et al. [37] and Ciardullo et al. [38] findings. We found underlying high blood pressure to be associated with COVID-19 mortality, which is in line with Li et al. [37] finding and is in contrast with Rawl et al. [39], Pei et al. [40], Sun et al. [31] and Ciardullo et al. [38] findings. Hospitalization in wards Was associated with COVID-19 related mortality, parallel with Chen et al. [41] findings, who found a relationship between ICU admission and mortality. The association between the need for mechanical ventilation and COVID-19 related mortality is in line with Chen et al. [41] and Zhou et al. [26] findings. The association between low oxygen saturation and low respiratory rate with mortality was in contrast with Sun et al. [31] findings.

In our previous study, anorexia, dry cough, anosmia and history of cancer were associated with COVID-19 related mortality [32], but in this study, we observed no relationship between mortality of COVID-19 and cancer, that may be due to different population of the study; two other provinces from one country. Anorexia showed a significant positive relationship with COVID-19 related mortality by Rawl et al. [39]. Regarding comorbidities, finding no significant association between cancer and COVID-19 related mortality is in line with Lee et al. [42] findings but is in contrast with Iftimie et al. [29], Mehta et al. [43], Dai et al. [44], Westblade et al. [45], Melo et al. [46], and Rüthrich et al. [47] findings. Different demographic features could explain this discrepancy. Finding no association between gender and COVID-19 related mortality is the same as Ruan et al. [35], Mehta et al. [43], Sun et al. [31]. Absence of association between fever and COVID-19 related mortality in our study is the same as our previous research [32], but it contrasts with the findings of Iftime et al. [29]. Myalgia, diarrhoea, nausea and vomiting were not predictors of mortality in our cohort, which contrast with Zhou et al. [26] findings.

The most important strength of this research is investigating impact of some new features on mortality rate of COVID-19 patients. Another important strength of this research is the large amount of the data used. However, there are some weaknesses. The patients are from a specific region. It is mentioned [48] that factors associated with mortality may differ in various regions. In addition, detailed clinical data on cardiovascular parameters were not available. Future research is needed investigating mortality of COVID-19 in heart disease patients.

## 5. Conclusion

In this research, we investigated the effect of some of the risk factors and symptoms of COVID-19 mortality rate for the first time. Our results show a significant association between mortality and risk factors like old age, headache, chest pain, respiratory distress, low respiratory rate, oxygen saturation less than 93%, need to a mechanical ventilator, having symptoms on CT, hospitalization in wards, time to infection, history of hypertension, neurological disorders, cardiovascular diseases, and having a risk factor or multiple risk factors. In contrast, there is no significant association between mortality and gender, fever, myalgia, dizziness, seizure, abdominal pain, nausea, vomiting, diarrhoea and anorexia. More studies are needed to confirm these findings. As future work, we will investigate the mortality rate of COVID-19 heart disease patients.

## Data Availability

The data that support the findings of this study are available on request from the corresponding author.

## Ethical approval

The study was approved by the Omid Hospital Ethics Committee.

---

**Table S2.**
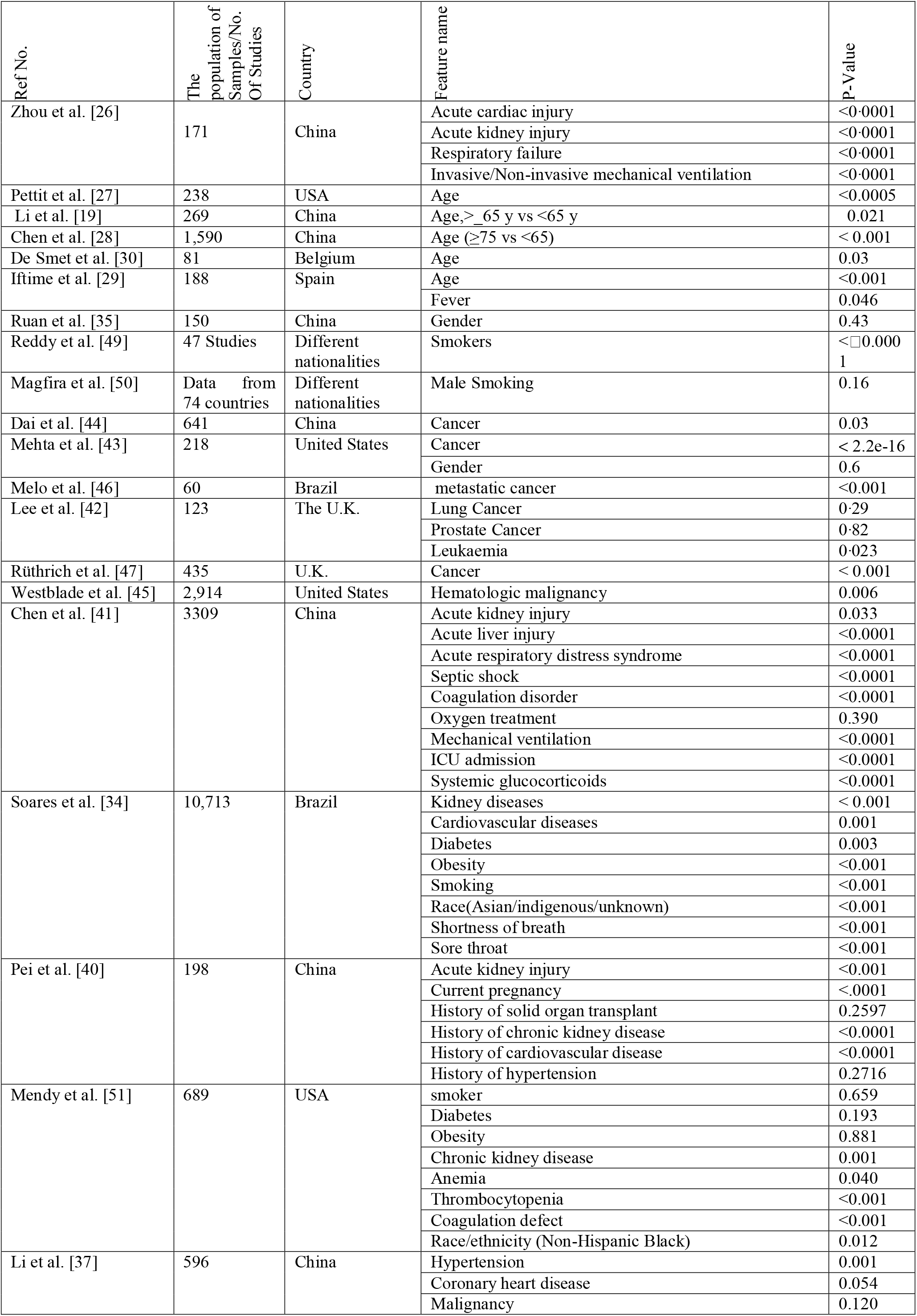
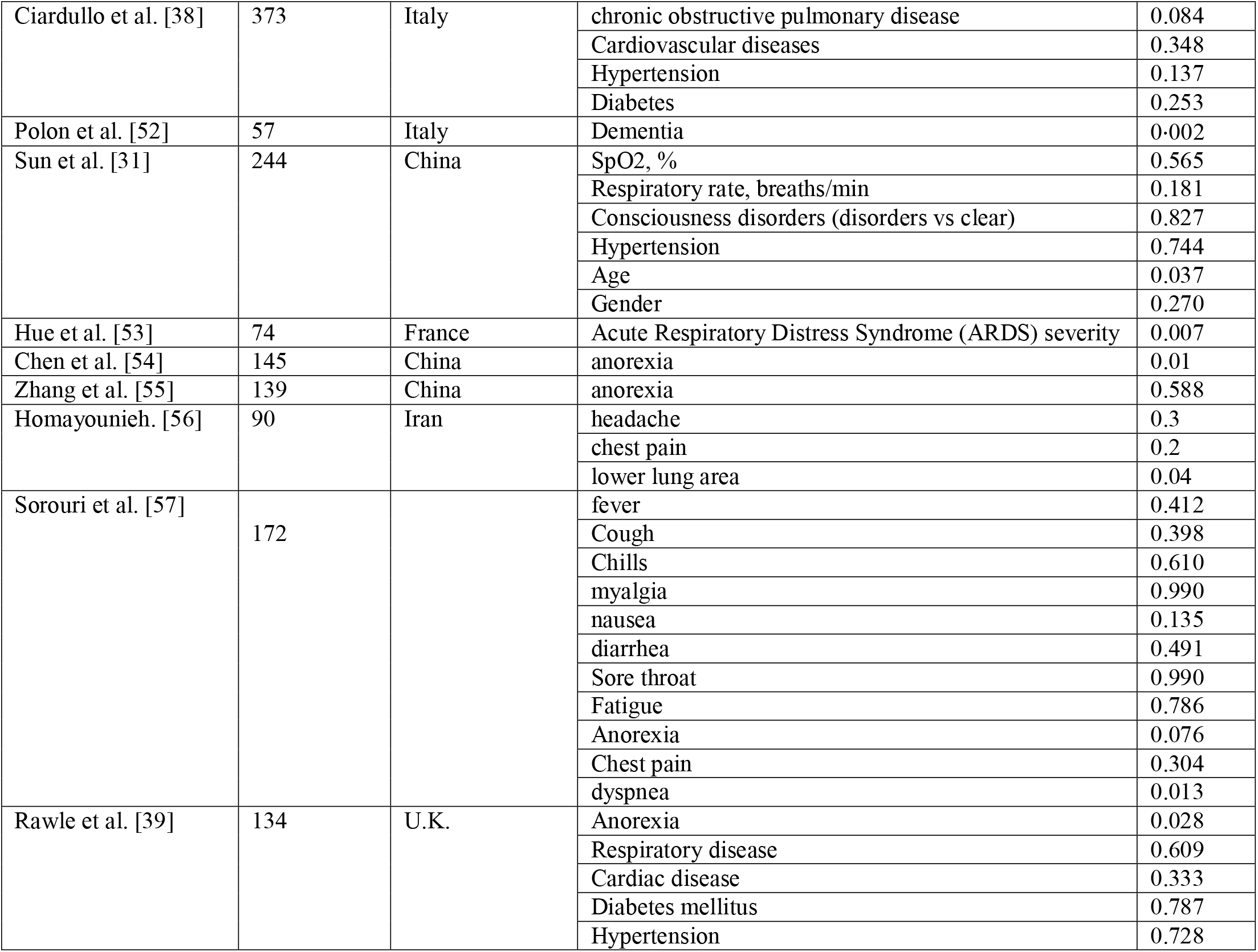
Some of the typical clinical characteristics of COVID-19 patients with mortality

| Ref No. | The population of Samples/No. Of Studies | Country | Feature name | P- Value |
| --- | --- | --- | --- | --- |
| Zhou et al. [26] | 171 | China | Acute cardiac injury | <0.0001 |
|  |  |  | Acute kidney injury | <0.0001 |
|  |  |  | Respiratory failure | <0.0001 |
|  |  |  | Invasive/Non-invasive mechanical ventilation | <0.0001 |
| Pettit et al. [27] | 238 | USA | Age | <0.0005 |
| Li et al. [19] | 269 | China | Age, >_65 y vs <65 y | 0.021 |
| Chen et al. [28] | 1,590 | China | Age (≥75 vs <65) | < 0.001 |
| De Smet et al. [30] | 81 | Belgium | Age | 0.03 |
| Iftime et al. [29] | 188 | Spain | Age | <0.001 |
|  |  |  | Fever | 0.046 |
| Ruan et al. [35] | 150 | China | Gender | 0.43 |
| Reddy et al. [49] | 47 Studies | Different nationalities | Smokers | <□0.0001 |
| Magfira et al. [50] | Data from 74 countries | Different nationalities | Male Smoking | 0.16 |
| Dai et al. [44] | 641 | China | Cancer | 0.03 |
| Mehta et al. [43] | 218 | United States | Cancer | < 2.2e-16 |
|  |  |  | Gender | 0.6 |
| Melo et al. [46] | 60 | Brazil | metastatic cancer | <0.001 |
| Lee et al. [42] | 123 | The U.K. | Lung Cancer | 0.29 |
|  |  |  | Prostate Cancer | 0.82 |
|  |  |  | Leukaemia | 0.023 |
| Rüthrich et al. [47] | 435 | U.K. | Cancer | < 0.001 |
| Westblade et al. [45] | 2,914 | United States | Hematologic malignancy | 0.006 |
| Chen et al. [41] | 3309 | China | Acute kidney injury | 0.033 |
|  |  |  | Acute liver injury | <0.0001 |
|  |  |  | Acute respiratory distress syndrome | <0.0001 |
|  |  |  | Septic shock | <0.0001 |
|  |  |  | Coagulation disorder | <0.0001 |
|  |  |  | Oxygen treatment | 0.390 |
|  |  |  | Mechanical ventilation | <0.0001 |
|  |  |  | ICU admission | <0.0001 |
|  |  |  | Systemic glucocorticoids | <0.0001 |
| Soares et al. [34] | 10,713 | Brazil | Kidney diseases | < 0.001 |
|  |  |  | Cardiovascular diseases | 0.001 |
|  |  |  | Diabetes | 0.003 |
|  |  |  | Obesity | <0.001 |
|  |  |  | Smoking | <0.001 |
|  |  |  | Race(Asian/indigenous/unknown) | <0.001 |
|  |  |  | Shortness of breath | <0.001 |
|  |  |  | Sore throat | <0.001 |
| Pei et al. [40] | 198 | China | Acute kidney injury | <0.001 |
|  |  |  | Current pregnancy | <.0001 |
|  |  |  | History of solid organ transplant | 0.2597 |
|  |  |  | History of chronic kidney disease | <0.0001 |
|  |  |  | History of cardiovascular disease | <0.0001 |
|  |  |  | History of hypertension | 0.2716 |
| Mendy et al. [51] | 689 | USA | smoker | 0.659 |
|  |  |  | Diabetes | 0.193 |
|  |  |  | Obesity | 0.881 |
|  |  |  | Chronic kidney disease | 0.001 |
|  |  |  | Anemia | 0.040 |
|  |  |  | Thrombocytopenia | <0.001 |
|  |  |  | Coagulation defect | <0.001 |
|  |  |  | Race/ethnicity (Non-Hispanic Black) | 0.012 |
| Li et al. [37] | 596 | China | Hypertension | 0.001 |
|  |  |  | Coronary heart disease | 0.054 |
|  |  |  | Malignancy | 0.120 |
| Ciardullo et al. [38] | 373 | Italy | chronic obstructive pulmonary disease | 0.084 |
|  |  |  | Cardiovascular diseases | 0.348 |
|  |  |  | Hypertension | 0.137 |
|  |  |  | Diabetes | 0.253 |
| Polon et al. [52] | 57 | Italy | Dementia | 0.002 |
| Sun et al. [31] | 244 | China | SpO2, % | 0.565 |
|  |  |  | Respiratory rate, breaths/min | 0.181 |
|  |  |  | Consciousness disorders (disorders vs clear) | 0.827 |
|  |  |  | Hypertension | 0.744 |
|  |  |  | Age | 0.037 |
|  |  |  | Gender | 0.270 |
| Hue et al. [53] | 74 | France | Acute Respiratory Distress Syndrome (ARDS) severity | 0.007 |
| Chen et al. [54] | 145 | China | anorexia | 0.01 |
| Zhang et al. [55] | 139 | China | anorexia | 0.588 |
| Homayounieh. [56] | 90 | Iran | headache | 0.3 |
|  |  |  | chest pain | 0.2 |
|  |  |  | lower lung area | 0.04 |
| Sorouri et al. [57] | 172 |  | fever | 0.412 |
|  |  |  | Cough | 0.398 |
|  |  |  | Chills | 0.610 |
|  |  |  | myalgia | 0.990 |
|  |  |  | nausea | 0.135 |
|  |  |  | diarrhea | 0.491 |
|  |  |  | Sore throat | 0.990 |
|  |  |  | Fatigue | 0.786 |
|  |  |  | Anorexia | 0.076 |
|  |  |  | Chest pain | 0.304 |
|  |  |  | dyspnea | 0.013 |
| Rawle et al. [39] | 134 | U.K. | Anorexia | 0.028 |
|  |  |  | Respiratory disease | 0.609 |
|  |  |  | Cardiac disease | 0.333 |
|  |  |  | Diabetes mellitus | 0.787 |
|  |  |  | Hypertension | 0.728 |

## Notes

### Competing Interest Statement

The authors have declared no competing interest.

### Funding Statement

there is no external funding

### Author Declarations

The study was approved by the Omid Hospital Ethics Committee.

